# Evaluation of the performance of a quantitative point-of-care CRP test

**DOI:** 10.1101/2022.05.20.22275259

**Authors:** JE Ellis, S MacLuskie, D Craig, L Lehane, G McInnes, J Harnett, G Cameron, P Moss, A Gray

## Abstract

**Introduction:** C-reactive protein (CRP) is an established acute-phase marker for infection and inflammation, which can help guide clinical decision-making in primary and secondary care. Many European guidelines recommend point-of-care (POC) CRP testing to improve antimicrobial stewardship in primary care. This performance evaluation study assessed the equivalence of the quantitative POC LumiraDx CRP Test compared to a laboratory-based reference method.

**Methods:** Method comparison, matrix equivalency, and precision were evaluated. Plasma samples from secondary care patients presenting with symptoms of infection or inflammation were analyzed centrally using the LumiraDx CRP Test and the reference test (Siemens CRP Extended Range for Dimension^®^ Clinical Chemistry System). The method comparison was conducted used Passing-Bablok regression analysis with prespecified criteria of r≥95 and a slope of 0.95–1.05. The REACT study (NCT05180110) evaluated the equivalence and precision of the testing modalities (fingerstick, venous blood, and plasma samples from the same secondary care patient) using Passing-Bablok regression analysis of the results of the POC LumiraDx CRP Test.

**Results:** In analysis of 320 plasma samples from 110 patients, the POC LumiraDx CRP Test demonstrated close agreement with the reference method, meeting the prespecified performance criteria (r=0.99, slope of 1.05, N=110). Paired replicate precision of the testing modalities was high, with mean %CV of 6.4 (plasma), 6.6 (capillary direct), and 8.1 (venous blood). Passing-Bablok regression showed matrix equivalency for all replicate pairs of the testing modalities, with r values across all sample types of 0.97–0.98.

**Conclusion:** The quantitative POC LumiraDx CRP Test showed very close agreement with the established laboratory-based test when using capillary blood, venous blood, or plasma. The use of capillary blood testing in particular is beneficial in both primary and secondary care, with this portable test system providing rapid quantitative results within 4 minutes, potentiating the ability to help guide clinical decision-making.

Data from two study collections, the NOVEL study and the REACT study with a trial registration: ClinicalTrials.gov identifier, NCT05180110, were used in this performance evaluation.

**Key summary points:**

- C-reactive protein (CRP) measurements are clinical markers for infection and inflammation, commonly used in primary and secondary care
- Point-of-care (POC) CRP testing can assist primary care clinicians in making an immediate decision as to whether to prescribe antibiotics while the patient is still at the clinic
- POC CRP testing that provides quantitative results near to the patient can be useful in emergency care assessment of patients and in hospital monitoring of antibiotic therapy
- The POC LumiraDx CRP Test has demonstrated quantitative results comparable to those obtained using a recognized laboratory system using plasma
- The POC LumiraDx CRP Test has also demonstrated matrix equivalence of capillary blood (both direct application and transfer tube), venous blood, and plasma

## Introduction

C-reactive protein (CRP) is an established clinical marker for infection and inflammation. It was discovered and named for its reactivity with the phosphocholine residues of C-polysaccharide, the teichoic acid of *Streptococcus pneumoniae* [1]. CRP is an annular, pentameric, acute-phase protein of hepatic origin, which can be found in blood plasma [1]. Its concentration in the circulation increases in response to infection and noninfectious inflammation, following increased secretion of inflammatory cytokines, particularly interleukin 6, by macrophages and T cells [2]. Conditions that affect CRP levels include rheumatoid arthritis, several cardiovascular diseases, and infection [2].

The average levels of CRP in the serum of a healthy Caucasian individual are approximately 0.8 mg/L, although this baseline can vary greatly depending on many factors that also influence plasma CRP levels, including heritability, sex, age, and body mass index [2, 3]. While there are no agreed standards for CRP levels, it is generally accepted that levels greater than 10 mg/L indicate inflammation or infection, marked elevation of more than 100 mg/L can signify acute bacterial and viral infection or major trauma, and levels greater than 500 mg/L indicate acute bacterial infection [4]. In the emergency department, as well as in primary care, CRP is measured in patients presenting with a variety of symptoms to aid detection and evaluation of infection, tissue injury, or inflammatory disorders [5], and to guide antibiotic treatment decisions [6].

To date, disease management for lower respiratory tract infections (LRTIs) in primary care has primarily been based on nonspecific clinical signs and symptoms, leading to overprescription of antibiotics [6]. The resulting increase in antimicrobial resistance has been identified as a major health concern by the European Commission and the World Health Organization [7]. Traditionally, diagnostic testing involved external laboratory analysis of venous blood (VB) draws, delaying initiation of therapy. Point-of-care (POC) CRP testing represents a rapid quantitative method using a simple fingerstick blood sample, which provides fast and accurate results at the consultation. The clinician can therefore make an immediate decision as to whether to prescribe antibiotics while the patient is still at the clinic. As such, the POC CRP test is a particularly useful tool in infection control and antimicrobial stewardship [6]. Guidelines in many European countries, in addition to guidance from the Joint Taskforce of the European Respiratory Society and European Society for Clinical Microbiology and Infectious Diseases outlining antibiotic prescription algorithms for LRTIs, recommend the use of POC CRP tests in primary care settings to limit over prescription of antibiotics [6, 8-11].

The POC LumiraDx CRP Test device uses microfluidic technology to detect and quantify CRP via a fluorescence immunoassay. The test uses magnetic nanoparticles coated with monoclonal anti-CRP antibodies and a fluorescent dye label. The assay can use fingerstick capillary blood, VB, or plasma samples (20 µL). When CRP is present, the resulting fluorescence is proportional to the CRP concentration in the sample. This study aimed to assess the equivalence, precision, and accuracy of the quantitative LumiraDx CRP Test when compared to the reference method, the Dimension^®^ Xpand^®^ Plus Integrated Chemistry System (Siemens; Munich, Germany), in patients with symptoms of infection, tissue injury, or inflammatory disorders.

## Methods

### Study design

Data from two study collections, the NOVEL study and the REACT study (NCT05180110), were used in this performance evaluation.

This prospective, observational study evaluated the performance of the LumiraDx CRP Test in samples from patients enrolled with symptoms of infection, tissue injury, or inflammatory disorders. The method comparison to evaluate the accuracy of the POC LumiraDx CRP Test compared to a reference device was carried out using samples from the NOVEL study. Standard venepuncture technique was performed by trained staff in five UK National Health Service (NHS) hospitals [12]. Lithium heparin anticoagulated whole blood samples were then sent to LumiraDx laboratories and processed to plasma for testing using the LumiraDx CRP Test and the reference device, the Dimension^®^ Xpand^®^ Plus Integrated Chemistry System. The NOVEL study complied with the Declaration of Helsinki (2013) and was approved by West of Scotland Research Ethics Committee 3 (REC number 15/WS/0176 and Integrated Research Application System ID: 179093).

The precision and accuracy of the LumiraDx CRP Test device using three different sample matrices was evaluated in patients presenting with symptoms of infection, tissue injury, or inflammatory disorders as part of the REACT study, which was performed in three UK NHS hospitals. Enrolled and consenting patients provided two venous whole blood samples (lithium heparin) and fingerstick capillary blood samples. Sample tubes of VB were collected from each participant using standard venepuncture technique performed by trained staff. A sample from one of the VB tubes was tested immediately using the LumiraDx CRP Test. The remainder of that tube and the second tube were processed to plasma and frozen for transport to the sponsor site (LumiraDx UK Ltd, Stirling, UK) for replicate testing and reference testing. A further four capillary blood samples were obtained from the participants. Two fingerstick samples (20 µL per test) were applied onto the test strip via direct application (DA), and two fingerstick samples (20 µL per test) were collected and applied to the test strip using a transfer tube (TT). The fingerstick samples were analyzed at the point of care using the LumiraDx CRP Test, following the manufacturer’s instructions for use [13]. The precision of the POC CRP assay was evaluated by way of mean paired replicate coefficient of variation (mprCV), which was presented as the mean percentage of the coefficient of variation (%CV). The REACT study received approval from the South-East Scotland Research Ethics Committee (REC 19/SS/0115) and the Health Research Authority. The study protocol (REC 19/SS/0115) complied with the Declaration of Helsinki (2013).

When testing samples using the LumiraDx CRP Test device, the time from preparing the test strip and collecting and applying the sample was approximately 1 minute, with a further 4 minutes required to obtain a test result. The number and type of error codes observed with the LumiraDx test device were recorded for both studies. The hematocrit (HCT) range for the LumiraDx CRP Test device was 25–55%.

### Study participants

Participants in both studies were secondary care patients who were aged 18 years or older. Written informed consent was obtained from all participants prior to enrolment. Full inclusion and exclusion criteria are outlined in the **Error! Reference source not found**..

### Statistical analysis

Required sample sizes were determined in line with Clinical and Laboratory Standards Institute (CLSI) guidelines (CLSI EP09c ED3:2018 and CLSI EP28 A3C:2010) [14, 15]. For the method comparison, plasma samples were measured using the LumiraDx CRP Test and Dimension^®^ Xpand^®^ Plus Integrated Chemistry System, and the average of two plasma replicates was taken. To assess method equivalency, a Passing-Bablok regression analysis was performed with pre-specified criteria of r≥95 and a slope of 0.95–1.05. The analysis of the paired replicate precision of the different testing modalities of the LumiraDx CRP Test was assessed using mprCV. A test with an analytical variability of ≤10% was considered to have high precision. Matrix equivalency of different testing modalities was assessed using Passing-Bablok regression, with the mean of two replicates analyzed across all measured sample types (capillary blood DA and TT, VB, and plasma). To demonstrate equivalence of methods, all confidence intervals of the slope must contain 1.0 [16]. The paired replicate precision analysis over time and sites was also assessed using mprCV, with ≤10% considered to indicate high precision. For the error rate analysis, the overall study error rate was calculated by analyzing all the raw data collected as part of the REACT and NOVEL studies.

Patient samples for the paired replicate precision analysis were excluded from the analysis if there were incomplete data sets for all three matrices owing to errors, missing replicates or results being outside the assay measuring range (<5 mg/L).

## Results

### Method comparison analysis

A total of 320 VB samples from the NOVEL study were collected between July 2019 and August 2020 from 110 patients ranging 18–88 years (median 61; mean 57) in age, 54.6% of whom were female. All participants in the NOVEL study were in hospital with suspected infection, injury, or inflammation, with a variety of presenting symptoms. The method comparison study was conducted with plasma samples across the CRP measuring range of 5.1–245.5 mg/L and using three test strip lots. Data analysis demonstrated close agreement of the LumiraDx CRP Test with the laboratory-based reference method, meeting the pre-specified performance criteria (r≥95 and a slope of 0.95–1.05) with r=0.99, a slope of 1.054, and an intercept of –0.66. The Passing-Bablok analysis showed the requirement of r^2^≥0.95 was met for all three test strip lots (0.98, 0.98, and 0.98) (**Table 1**).

**Table 1:**
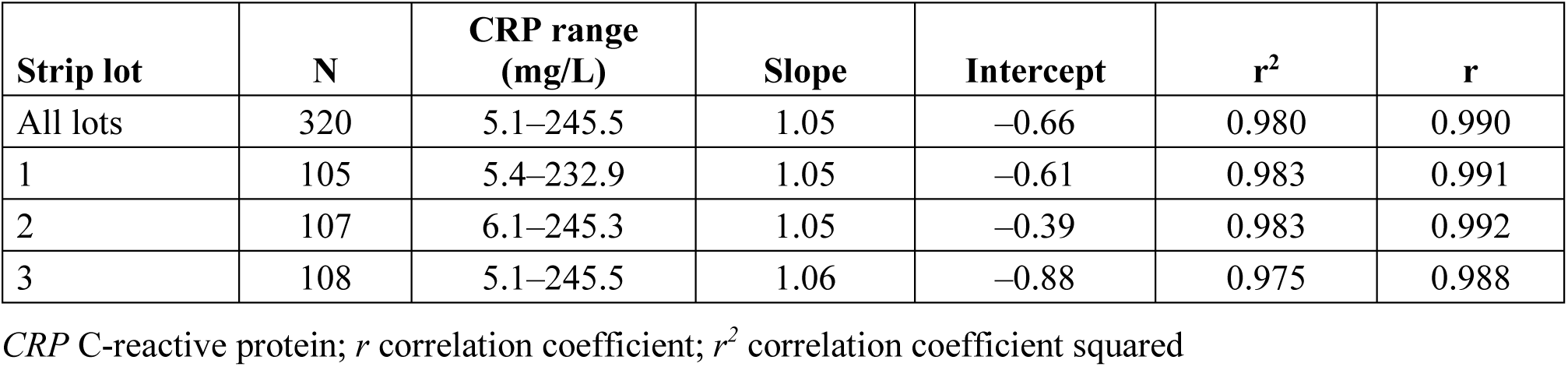
The Passing-Bablok regression results displayed as LumiraDx CRP plasma test results versus Dimension^®^ Xpand^®^ Plus Integrated Chemistry System plasma test results

| Strip lot | N | CRP range (mg/L) | Slope | Intercept | r<sup>2</sup> | r |
| --- | --- | --- | --- | --- | --- | --- |
| All lots | 320 | 5.1–245.5 | 1.05 | –0.66 | 0.980 | 0.990 |
| 1 | 105 | 5.4–232.9 | 1.05 | –0.61 | 0.983 | 0.991 |
| 2 | 107 | 6.1–245.3 | 1.05 | –0.39 | 0.983 | 0.992 |
| 3 | 108 | 5.1–245.5 | 1.06 | –0.88 | 0.975 | 0.988 |

CRP C-reactive protein; *r* correlation coefficient; *r*<sup>2</sup> correlation coefficient squared

### Paired replicate precision and matrix equivalency

As part of the REACT study, samples from 57 participants were collected between November 25^th^, 2021 and December 8^th^, 2021. The age range of participants was 18–84 years (median 55; mean 54) and 63% were female. Paired replicate precision and matrix equivalency analysis was conducted by POC operators on 44 participants with complete sample sets, where fingerstick blood (DA and TT), VB, and plasma results were compared in duplicate. Paired replicate precision was assessed across a CRP measuring range of 19.9–185.4 mg/L for each sample type. The mean %CV for the three sample types was 6.4 for plasma, 6.6 for capillary DA, and 8.1 for VB (**Table 2**). Matrix equivalency analysis for the different test modalities using Passing-Bablok regression resulted in r values of 0.97–0.98 across all sample types, demonstrating matrix equivalency for all pairs (**Table 3**).

**Table 2:**
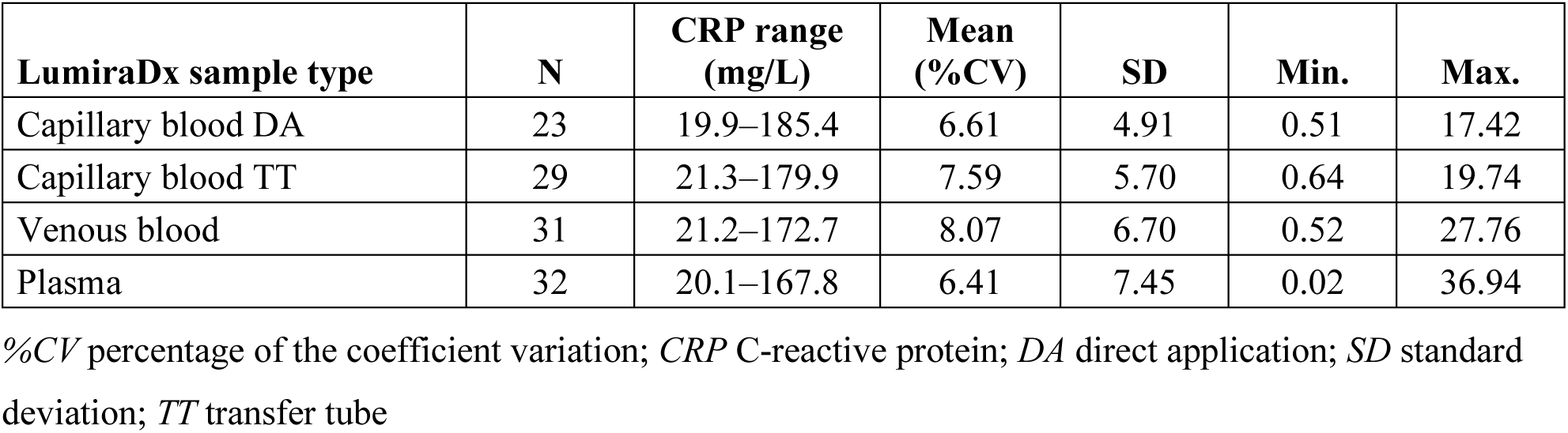
Paired replicate precision analysis of the LumiraDx CRP assay

**Table 3:** Passing-Bablok regression analysis assessing matrix equivalency for the different testing modalitiess

| LumiraDx sample type | N | CRP range (mg/L) | Slope | Intercept | r |
| --- | --- | --- | --- | --- | --- |
| Plasma vs venous blood | 43 | 5.2–69.6 | 1.05 | –0.79 | 0.981 |
| Plasma vs capillary blood TT | 44 | 5.2–169.6 | 0.93 | 0.90 | 0.977 |
| Plasma vs capillary blood DA | 44 | 5.2–169.6 | 0.98 | –0.29 | 0.974 |
| Venous vs capillary blood TT | 44 | 5.7–198.5 | 0.95 | 0.73 | 0.983 |
| Venous vs capillary blood DA | 43 | 5.7–198.5 | 0.98 | 0.09 | 0.974 |
| Capillary blood TT vs DA | 44 | 5.1–221.2 | 1.06 | –1.21 | 0.982 |

CRP C-reactive protein; DA direct application; r correlation coefficient; TT transfer tube

### Error rate analysis

The error rate analysis showed an overall study error rate of 2.9% for the combined REACT and NOVEL studies. Of the 31 test strip errors, 29 resulted from the REACT venous and capillary blood testing, while the REACT plasma testing resulted in no errors. The remaining two test strip errors occurred as part of the NOVEL study. The most common test strip error in the REACT study was recorded as Door Close Timeout (n=9). This error was corrected with product training. The second most common error (n=7) resulted from unsuitability of samples owing to the HCT parameters being out of range.

## Discussion

The performance evaluation, in which the accuracy of the quantitative LumiraDx CRP Test was compared to the laboratory-based reference method, demonstrated equivalence between methods in patients with symptoms of infection, tissue injury, and inflammatory disorders. The accuracy of the LumiraDx CRP Test was established across a range of CRP concentrations, and the matrix equivalency was proven for whole blood fingerstick samples (DA or TT), VB, and plasma. The precision analysis of the assay showed high precision across the different sample types. Furthermore, the LumiraDx CRP Test showed a low overall error rate of 2.9%.

POC CRP tests are used in primary care in many countries [6]. Many guidelines in Europe recommend using a POC test for patients presenting with symptoms of LRTI and acute exacerbations of chronic obstructive pulmonary disease (COPD) in primary care, to guide the physician’s antibiotic prescription decision [6]. In fact, recommendations around the use of POC CRP tests for respiratory infections in primary care settings have been included in guidelines in Norway, Sweden, the Netherlands, Germany, Switzerland, the Czech Republic, Estonia and the UK [6, 10]. The accuracy of POC CRP tests compared to laboratory testing such as chest X-rays has also been demonstrated in diagnostic studies of community-acquired pneumonia [17]. An association has been drawn with reduced levels of antibiotic prescribing in the countries implementing POC CRP tests [6]. In countries where these tests are not yet consistently implemented in practice, it could be a result of the need for better guidance and training, and resource constraints [6, 18, 19].

A recent systematic literature review and meta-analysis examined studies of patients who presented at primary care with respiratory tract infections (RTIs) which included one of four types of POC CRP test: NycoCard (II) CRP readers (Abbott, Chicago, IL, USA), QuickRead go CRP (Aidan, Espoo, Finland), Afinion CRP (Abbott), or CRP multichannel analyzer. All the studies compared POC CRP tests with a control intervention consisting of usual care, and the results demonstrated that using POC CRP tests to guide antibiotic prescribing for (lower and upper) RTIs in primary care reduced antibiotic prescribing, especially when cut-off guidance was provided. The resulting reduction in antibiotic prescribing appeared to increase the re-consultation rate but did not affect clinical recovery, resolution of symptoms, or hospital admissions [20]. POC CRP testing is also of value for patients with exacerbations of COPD, which is a patient group that is associated with high antibiotic use. In a multicenter, open-label, randomized controlled trial of patients at primary care presenting with exacerbations of COPD, there was significantly lower antibiotic among the patients who received CRP-guided prescribing of antibiotics with no evidence of harm compared with patients who received usual care alone without CRP guidance [18].

The majority of studies examining the effect of POC CRP tests on patients with LRTIs that present at primary care have found a reduction in antibiotic prescribing compared with usual care, and physicians and patients alike report high acceptability of the use of POC CRP tests [6]. The POC LumiraDx CRP Test has the potential to guide primary care physicians in the clinical decision pathway and thereby reduce rates of unnecessary antibiotic prescription. Furthermore, the LumiraDx CRP Test can be performed in a few minutes, requiring a finger stick for sampling rather than VB collection, and allows the physician to make the appropriate clinical decision during the consultation with the patient, thereby enabling more rapid implementation of the indicated treatment. Removing the need for samples to be sent for external laboratory analysis saves time and resources and reduces the need for follow-up appointments. Qualitative studies suggest that it may also increase the patient’s trust in the treatment decision, potentially leading to improved therapy adherence [21, 22].

The limitations of this study include the use of two different patient cohorts when performing the method comparison and matrix equivalency analyses, which was due to restrictions resulting from the coronavirus disease 2019 pandemic. This study was performed in secondary care by trained research teams and compared against a laboratory-based CRP measurement system. However, the clinical performance of the test would not be expected to be affected by this setting. As the primary objective of this study was to assess comparative performance against an established laboratory instrument, the clinical impact of the system, including with regards to reducing antibiotic prescription, was not directly addressed. Usability of the LumiraDx CRP Test at primary care sites was not evaluated in this study. However, the LumiraDx Platform has previously been evaluated on the basis of use of D-Dimer tests at primary care sites, in which capillary blood, VB, and plasma were also sample types [23]. The LumiraDx system was demonstrated to be easy to use. As the test workflow is the same between the LumiraDx D-Dimer and CRP tests, the usability is expected to be the same [23].

## Conclusion

The high accuracy and precision, low error rate, and choice of different testing modalities demonstrate how the LumiraDx CRP Test is a good alternative to the established laboratory-based reference method using plasma samples when performed by research teams in secondary care. It provides the benefit of delivering a quantitative result from a fingerstick sample in 4 minutes at the point of care. The evidence suggests the potential of the LumiraDx CRP Test in guiding primary care physicians in their decision to prescribe antibiotics during patient consultations.

## Data Availability

All data produced in the present study are available upon reasonable request to the authors.

## Authorship

All named authors meet the International Committee of Medical Journal Editors criteria for authorship for this article, take responsibility for the integrity of the work as a whole and have given their approval for this version to be published.

## Author contributions

Jayne Ellis, Sarah MacLuskie, David Craig, Lucy Lehane, and Graeme McInnes were responsible for the concept and design of the study, as well as drafting the manuscript. Sarah MacLuskie, David Craig, and Graeme McInnes generated the data. Jayne Ellis, Sarah MacLuskie, David Craig, Lucy Lehane, Graeme McInnes, James Harnett, Gregor Cameron, Phil Moss, and Alasdair Gray all contributed to the evaluation of the clinical data and critically reviewed the manuscript.

## Medical writing, editorial, and other assistance

The authors acknowledge Anne-Marie Quirke and Kathrin Schulze-Schweifing, of integrated medhealth communication (imc), for medical writing support.

## Compliance with ethics guidelines

The NOVEL study complied with the Declaration of Helsinki (2013) and was approved by West of Scotland Research Ethics Committee 3 (REC number 15/WS/0176 and Integrated Research Application System ID: 179093). The REACT study received approval from the South-East Scotland Research Ethics Committee (REC 19/SS/0115) and the Health Research Authority. The study protocol (REC 19/SS/0115) complied with the Declaration of Helsinki (2013). All participants provided informed consent prior to participation.

## Data availability

The datasets generated during and/or analyzed during the current study are available from the corresponding author on reasonable request.

## Thanking patient participants

The authors would like to extend their gratitude to the study participants for their involvement in the NOVEL and REACT studies

