## supplemental file for "Evaluation of the performance of a quantitative point-of-care CRP test"

### Supplementary files

#### REACT study inclusion and exclusion criteria

Participants included patients who presented in emergency departments, surgical patients, patients presenting at acute medical units, or in outpatient clinics with symptoms indicative of infection, injury, or inflammatory disorders. These included but were not limited to: respiratory tract infection (upper and lower), rheumatoid arthritis, lupus, burns, trauma, inflammatory bowel disease.

Patients were excluded from participation in the REACT study if they were under 18 years of age currently receiving, or had received within the past 30 days of the study visit, an experimental biologic or drug (including either treatment or therapy); had skin lesions or conditions that would preclude a fingerstick and/or a VB draw; had previously participated in this study; were critically ill or required a time-critical intervention; were receiving end-of-life or palliative care; suffered from myeloma, monoclonal gammopathy or extreme lipemia; or were deemed medically unfit to participate.

#### NOVEL study inclusion and exclusion criteria

Participants included patients attending hospital with symptoms indicative of heart failure, acute coronary syndrome, kidney failure, thromboembolic events, or inflammatory disorders.

Patients were excluded from the NOVEL study if they were under 18 years of age or belonged to vulnerable populations deemed inappropriate for the study by the site principal investigator.
